# Genome-wide association study of extrapulmonary traits in the context of COPD

**DOI:** 10.64898/2026.02.23.26346864

**Authors:** Rui Marçalo, Guilherme Rodrigues, Cíntia Dias, Ana Sofia Grave, Raquel Vilar Marinho, Miguel Pinheiro, Sonya Neto, Stephanie Holum, Ana Rita Guimarães, Sofia Lucília Marques, Paula Simão, Vitória Martins, Lília Andrade, Maria Aurora Mendes, Manuel António da Silva Santos, Rosa Faner, Sandra Casas-Recasens, Borja García-Cosio, Àlvar Agustí, Corry-Anke Brandsma, Maarten van den Berge, Alda Marques, Gabriela Ribeiro Moura

## Abstract

Functional capacity, muscle strength, and patient-reported outcome measures are important indicators of health. In chronic obstructive pulmonary disease (COPD), these traits are often impaired beyond normal age-related decline. Substantial variability exists in both COPD and healthy populations, the biological basis of which remains poorly understood. Given the known contribution of genetics to complex traits, genetic factors may partly explain this variability. This study aimed to identify genetic variants associated with measures used to characterise extrapulmonary traits in COPD.

Genome-wide association studies were conducted on the Lab3R-ESSUA cohort for the 6-minute walk test (6MWT), the 1-minute sit-to-stand test (1-min STS), the quadriceps maximal voluntary contraction (QMVC), the handgrip muscle strength, and the chronic airways assessment test (CAAT), adjusting for age, sex, body mass index, pack-years and ancestry. Variants with P<1E^-05^ were selected for replication in the EARLYCOPD cohort, and effects compared between COPD and healthy populations (two-way ANOVA).

A total of 639 participants (364 people with COPD, 275 healthy; 75% male, median age 67 years; BMI of 27 Kg/m^2^; 10 pack-years) were included. Significant variants were identified for the 6MWT (rs1108983:G, β=-186.5m, P=4.8E^-08^), the 1-min STS (rs5889103:GTT, β=4.2reps, P=4.8E^-08^), the Handgrip (rs67352743:A, β=-4.4Kg, P=2.8E^-08^), and for the CAAT (rs11747040:C, β=4.4points, P=4.0E^-09^; rs11041680:A, β=-2.6points, P=2.5E^-08^). Effects were independent of COPD diagnosis. Replication in EARLYCOPD (n=282) confirmed one SNP for 6MWT and three for CAAT.

These findings highlight genetic contributions to functional capacity, muscle strength, and disease burden. COPD-related impairments appear to build on pre-existing genetic predisposition, contributing to disease heterogeneity.

## Introduction

Chronic obstructive pulmonary disease (COPD) is a heterogenous and complex condition characterized by persistent airflow limitation and respiratory symptoms^1,2^. These are driven by structural and functional alterations at the lungs, such as emphysema, and airways, such as chronic bronchitis^3^. Historically, research on COPD has primarily focused on these pulmonary traits^4–9^. However, growing recognition of COPD as a multifactorial disease has shifted attention towards a broader spectrum of traits that extends beyond the lungs^10–12^. These extrapulmonary traits provide a more comprehensive understanding of patients’ health status, often surpassing pulmonary traits alone in clinical relevance^13–16^. Functional capacity and muscle strength are key components of functional status, reflecting an individual’s ability to perform daily tasks and fulfil their social roles^15–18^. In disease contexts such as COPD, these traits are commonly impaired and part of a health-decaying cascade of events that culminate in loss of independence and the need of a caretaker, in many cases a family member^17^. This is reflected in patients’ perception of disease burden, which plays a crucial role in assessing the true impact of COPD on daily life^19,20^. Although commonly impaired in disease contexts (e.g., COPD), patients exhibit varying degrees of impairments across different traits, that cannot be fully explained by the disease alone^21^. Moreover, these traits also deteriorate naturally with aging, emphasizing their importance not only in specific diseases but also to the general population^22,23^.

These extrapulmonary traits are influenced by a combination of genetic, environmental, and clinical factors^24–27^. However, few studies have explored the influence of genetics on extrapulmonary traits that directly affect patients’ daily lives, and often lack replication due to the limited number of cohorts with similar available data^6,24–26^. Unravelling the genetic background of these traits can enhance our understanding of systemic health, clarify the heterogeneity observed in COPD, and offer valuable insights into potential preventive measures and therapeutic interventions.

This study aims to identify genetic variants associated with functional capacity, muscle strength and impact of disease, while exploring the biological pathways underlying these associations through genome-wide association studies (GWASs) in a Portuguese cohort composed of people with COPD and healthy individuals.

## Methods

### Ethics approval

A cross-sectional secondary analysis was conducted using data collected from 2017 to 2023, in GENIAL (PTDC/DTP-PIC/2284/2014), PRIME (PTDC/SAU-SER/28806/2017; POCI-01-0145-FEDER-028806), 3R (SAICT-POL/23926/2016; POCI-01-0145-FEDER-016701), and CENTR(AR) (POISE-03-4639-FSE-000597) projects. Approval for this study was obtained from the Ethics Committees of the Administração Regional de Saúde do Centro, I.P. (3NOV’2016:64/2016), Centro Hospitalar do Baixo Vouga (22MAR’2017:777638),), Hospital Distrital da Figueira da Foz (18JUL’2017), Unidade Local de Saúde de Matosinhos (17FEB’2017:10/CE/JAS) and Centro Hospitalar do Médio Ave (27AUG’2018 and from the National Data Protection Commission (8828/2016). Informed written consents were collected from each participant before any data collection. This study is reported according to the Strengthening the Reporting of Observational Studies in Epidemiology (STROBE) guidelines^28^.

### Recruitment

The Lab3R-ESSUA Cohort is composed of people with COPD and healthy individuals. Recruitment of those with COPD was performed by physicians, from hospitals and primary healthcare centres of the Centre Region of Portugal during routine medical appointments. Individuals with COPD were deemed eligible if presenting a diagnosis of COPD according to GOLD criteria^1,2^, clinically stable in the previous month, and able to provide informed consent. Healthy individuals were recruited from routine appointments in primary health care centres and senior universities. They were deemed eligible if over 18 years old, considered “healthy” by the general family practitioner, i.e., with the most prevalent age-related conditions (e.g., controlled arterial hypertension, dyslipidaemia, diabetes) to ensure maximum representativeness from community-dwelling people^29^. Exclusion criteria for all participants included the presence of a diagnosed respiratory disease (excluding COPD), history of neoplasia, severe cardiac/musculoskeletal/neuromuscular diseases, signs of cognitive impairment, or immune disease that would interfere in data collection or interpretation.

### Data collection

Sociodemographic (age, sex), anthropometric (height, weight), and clinical data (smoking, muscle strength, functional capacity, disease impact) were collected upon enrolment in the project with a structured protocol. Quadriceps maximal voluntary contraction (QMVC) was measured with a handheld dynamometer (microFET2, Hoggan Health, Salt Lake City, Utah), in kilogram-force (Kg.F), during 6 seconds with resistance applied to the anterior tibia (patient in a seated position)^30^. Handgrip muscle strength (Handgrip) was measured with a dynamometer (Baseline® Hydraulic Hand Dynamometer), in kilogram (kg), during 3 seconds with the patient seated with the wrist neutral and the elbow flexed to 90 degrees^31^. All muscle strength measurements were taken at the dominant side. After two repetitions without resistance (familiarisation purposes), the best out of three acceptable and reproducible manoeuvres (less than 10% variation) was considered^25,26^. The 6-minute walk test (6MWT) and the 1-minute sit-to-stand test (1-min STS) were performed twice and the best performance was registered to assess functional exercise capacity^32,33^. The percentage predicted values for the 6MWT, the 1-min STS, the QMVC and the Handgrip were calculated based on reference equations^34–37^. The impact of the disease was measured with the Chronic Airways Assessment Test (CAAT)^38^. The CAAT scores range from 0 to 40, with higher scores indicating higher disease impact, and a cut-off of 10 points used in the GOLD ABE assessment tool to classify patients with high symptom burden, according to the GOLD guidelines^20^.

### Genotyping, quality control and imputation

Saliva samples were collected from all individuals and DNA extracted using QIAamp DNA Blood Mini Kit (QIAGEN, Hilden, Germany), following manufacturer’s protocol. Genotyping of DNA was performed using the Infinitum Global Screening Array-24 v1.0. Quality control of both samples and SNPs was performed using GenomeStudio 2.0 and PLINK 1.9 software, according to standard protocols^39,40^. Briefly, samples with genotyping rate below 95%, divergent ancestry from the study population, sex discrepancies, a heterozygosity rate higher than predicted, or unreported relatedness to another study participant were excluded from the analysis. In addition, SNPs with a genotyping rate below 95%, deviation from Hardy-\Weinberg equilibrium, or a minor allele frequency below 5% were also excluded. The Michigan imputation server was used for SNP imputation^41^, followed by SNP quality control again. In total, 639 individuals and 7.459.320 SNPs passed quality control.

### Genome-wide association study and SNP functional enrichment analysis

A logistic regression adjusted for age, sex, body mass index (BMI), smoking load (pack-years) and the first four principal components to adjust for genetic ancestry was performed for the 6MWT, the 1-min STS, the QMVC, the Handgrip and the CAAT, in order to identify genetic variants associated with these extrapulmonary traits. An additive model was assumed, and significance set at P=5E^-8^ and P=1E^-5^ for significant and suggestive findings, respectively^42,43^. Analysis was carried out using PLINK 1.9 software^44^. FUMA_GWAS_ was used for mapping and functional annotation of the identified SNPs^34^. Identified genes were assessed for pathway enrichment using web-based gene set analysis toolkit and snpXplorer^45,46^.

### SNP x Diagnosis interaction

The effects of significant SNPs were compared between people with COPD and healthy individuals to assess whether the SNPs were specific to the trait rather than being influenced by COPD. A two-way ANOVA was conducted for each trait, having the trait as dependent variable and the interaction term SNP x diagnosis as independent variable. If ANOVAs’ residuals were not normally distributed, a non-parametric alternative (aligned rank transformation) was applied instead. Significance was set at P=0.05 for the interaction term. Analysis was carried using the ARTool package and built-in functions of R software (version 4.2.1.)^47,48^.

### Replication

Replication of the SNPs associated with the 6MWT and the CAAT was performed using data from the EARLYCOPD cohort (early COPD and CEPA studies)^49,50^. Briefly, the EARLYCOPD is a multicentre, prospective case-control cohort of people with COPD that aimed to characterize early-stage COPD by collecting multidimensional data on demographics, lung function, exercise capacity, quality of life, biomarkers, and comorbidities. The 6MWT and the CAAT were modelled with a logistic regression, that included the SNP being replicated and the covariables age, sex, smoking status and the first 4 principal components. Built-in functions from R software (version 4.2.1.) were used^47^. Significance was set at P=0.05.

Replication of the SNPs associated with the Handgrip and the QMVC was not possible due to inaccessibility to genotyped cohorts characterized for these variables. Alternatively, our cohort was used to validate previously reported genetic variants associated with the QMVC and the Handgrip. SNPs were identified from the GWAS Catalog database^51^ under the search terms “Handgrip” and “lower body strength measurement”. Selected SNPs were retrieved from the GWASs of the Lab3R-ESSUA Cohort and compared to the original study that reported their association with either the QMVC or the Handgrip. Significance was set at P=0.05.

No genetic variants have been previously reported as associated with the 1-min STS nor a cohort with genotyped participants characterized for the 1-min STS was available for replication.

## Results

### Cohort characterisation

Of the initial 639 participants (COPD=364 and healthy=275) with available genotypic data, 227 had data for the 6MWT, 598 had data for the 1-min STS, 627 had data for the QMVC, 567 had data for the Handgrip, and 610 had data for the CAAT (Figure 1). Participants had a median age of 67 [60-73] years, were predominantly male (75.1%), overweight (median BMI of 26.9 [24.2-30.3] Kg/m^2^) and presented a median smoking load of 10 [0-43.3] pack-years. The description of the cohort can be found in table 1.

**Table 1.**
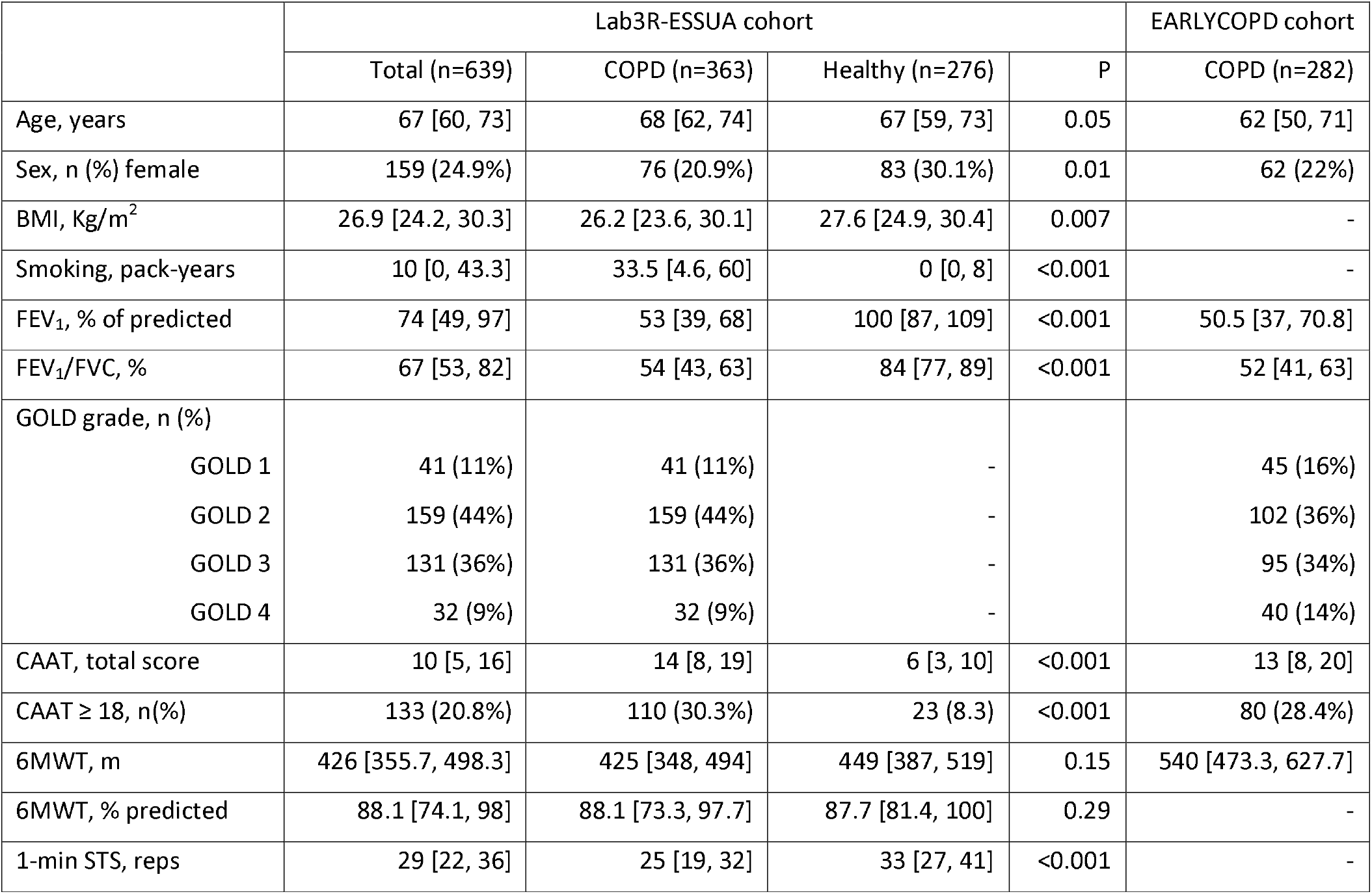
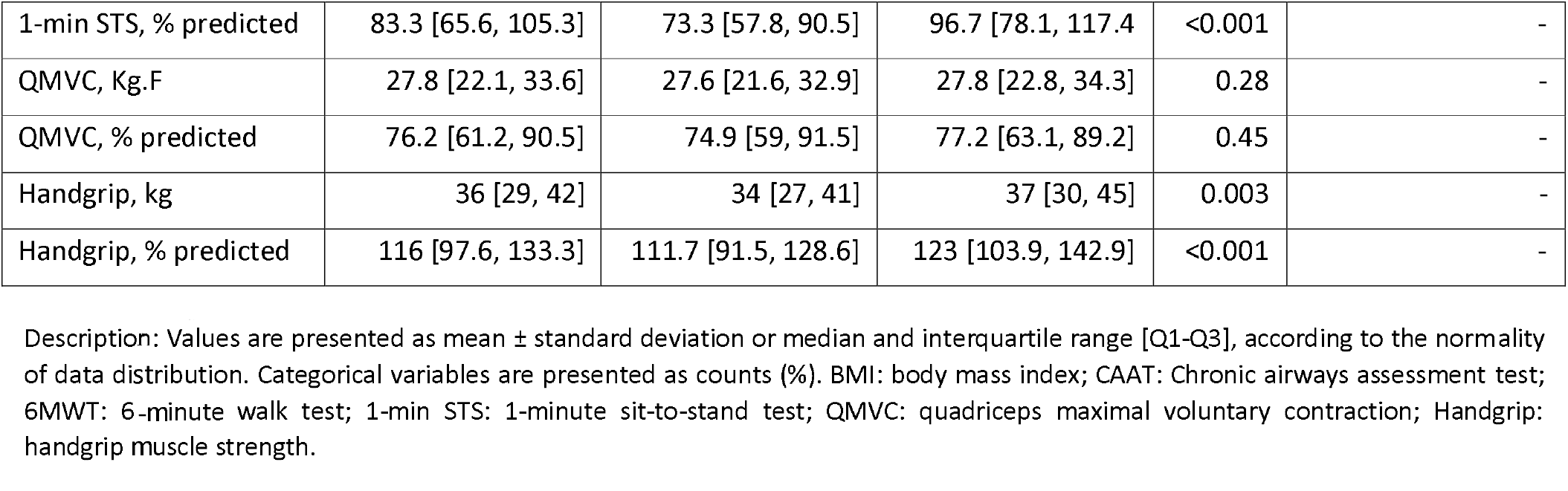
Sociodemographic, anthropometric and clinical characteristics of the Lab3R-ESSUA Cohort (n=639) and the replication cohort EARLY COPD (n=282).

**Figure 1.**
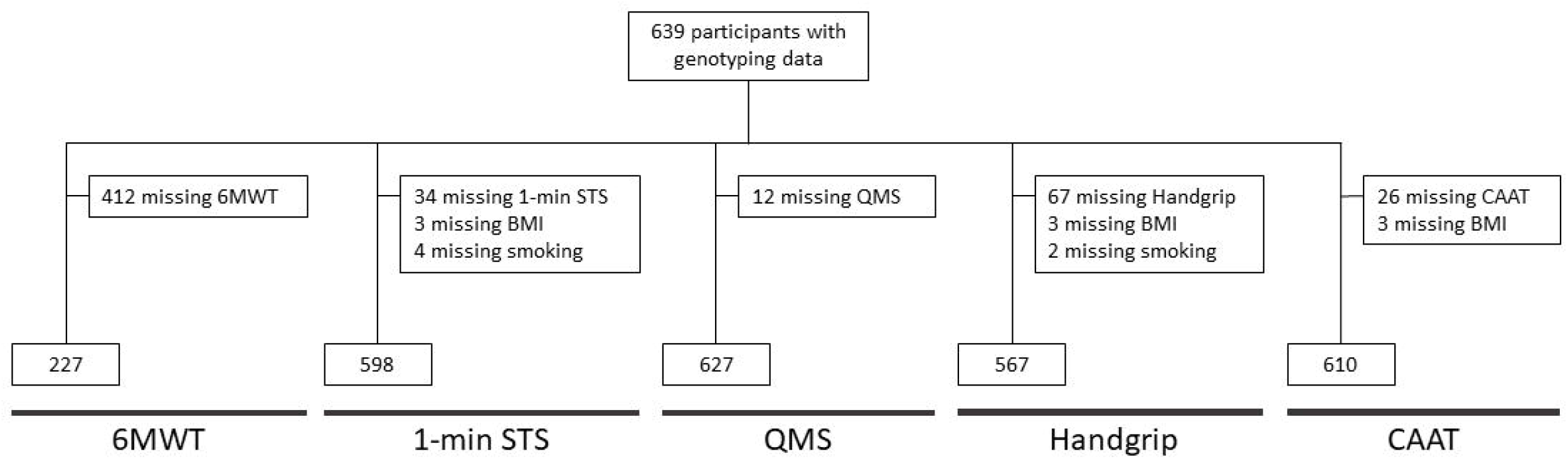

### Genetic association analysis

#### 1. 6-minute walk test

We found 115 SNPs suggestively associated (P<1E^-05^) with the total distance walked in the 6MWT and one SNP that reached genome-wide significance (rs1108983:G, β (SE)= -186.5 (30.9) meters, P=4.8E^-08^) (Figure 2A). People with A/A genotype for rs1108983 averaged 447.1±88.6 meters in the 6MWT, whereas people with A/G genotype walked on average 241.7±121.1 meters over the same duration. Rs1108983 was mapped to the gene Tumor Necrosis Factor Superfamily Member 8 (*TNFSF8*) in chromosome 9. The 115 suggestive SNPs were mapped to 49 different genes (Supplementary table 1). The identified genes showed no significant enrichment for any pathway (Supplementary figure 1A).

**Figure 2.**
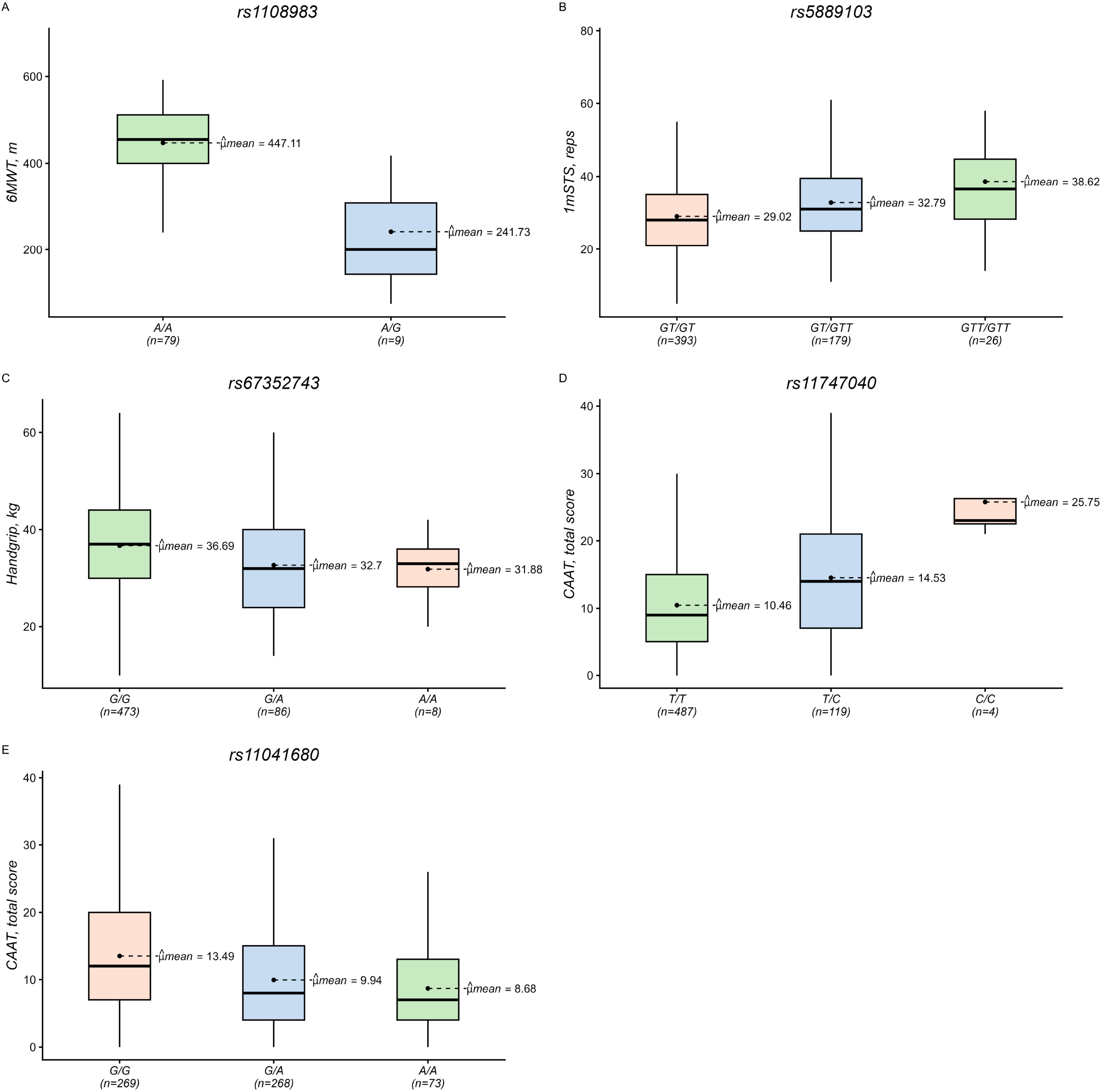

#### 2. 1-minute sit-to-stand test

The GWAS on the 1-min STS unravelled 96 SNPs suggestively associated (P<1E^-05^) with the phenotype. Additionally, one SNP reached genome-wide significance (rs5889103:GTT, β (SE)= 4.2 (0.7) repetitions, P= 4.8E^-08^). The presence of the minor allele was associated with an increase in the number of repetitions performed during the 1-min STS test (GT/GT: 29±10.9 reps; GT/GTT: 32.8±11.8 reps; GTT/GTT: 38.6±14.2 reps) (Figure 2B). Rs5889103 is an intergenic insertion-deletion (indel) variant on chromosome 8. The remaining 96 SNPs were mapped to 27 genes and GSEA unravelled 8 pathways, mainly involved in glycosylation, ion channels activation and extracellular matrix remodelling, where a significant enrichment was observed (Supplementary table 1 and Supplementary figure 1B).

#### 3. Quadriceps maximal voluntary contraction

We found no genome-wide significant SNPs associated with the QMVC. However, 123 SNPs emerged as suggestively associated with QMVC, which mapped to 73 genes (Supplementary table 1). GSEA revealed a significant enrichment in genes associated with Williams-Beuren syndrome (Supplementary figure 1C).

#### 4. Handgrip muscle strength

We identified one SNP, rs67352743, significantly associated with Handgrip (rs67352743:A, β (SE)= -4.4 (0.8) Kg, P= 2.8E^-08^) and 82 SNPs suggestively associated. The A allele of rs67352743 was associated with reduced handgrip muscle strength (G/G: 36.7±10.1 Kg; G/A: 32.7±10.7 Kg; A/A: 31.9±7 Kg), with no differences between having one or two A alleles (Figure 2C). Rs67352743 is an intergenic variant located in chromosome 1. In addition, 82 SNPs in 39 genes were suggestively associated with Handgrip (Supplementary table 1). GSEA identified enrichment in the signalling pathways of *L1CAM* and *CHL1* (Supplementary figure 1D).

#### 5. Chronic airway assessment test

The GWAS on the CAAT unravelled 133 SNPs associated with the phenotype. Of the 133 SNPs identified, 11 SNPs across two loci reached genome-wide significance (top hits per loci: rs11747040:C, β (SE)= 4.4 (0.7) points, P=4.0E^-09^; and rs11041680:A, β (SE)= -2.6 (0.5) points, P=2.5E^-08^) (Figure 2D and 2E). The two minor alleles showed opposite effects on CAAT total score. The increase in C alleles of rs11747040 was associated with a higher CAAT score (T/T: 10.5±7.5 points; T/C: 14.5±9 points; C/C: 25.8±6.9 points) (Figure 2D), whereas the increase in A alleles of rs11041680 was associated with a lower CAAT score (G/G: 13.5±8.5 points; G/A: 9.9±7.3 points; 8.7±6.8 points) (Figure 2E). Rs11747040 and rs11041680 are intronic variants of Corticotropin Releasing Hormone Binding Protein (*CRHBP*) in chromosome 5, and Olfactory Receptor Family 10 Subfamily A Member 3 (*OR10A3*) in chromosome 11, respectively. The suggestively associated SNPs were mapped to 33 genes (Supplementary table 1), with no enriched pathways emerging from the GSEA (Supplementary figure 1E).

### SNP x Diagnosis interaction

The effect of the significant SNPs was not different in people with COPD compared to healthy individuals (1-min STS[rs5889103]: p=0.96; Handgrip[rs67352743]: p=0.97; CAAT[rs11747040]: p=0.72; CAAT[rs11041680]: p=0.96) (Supplementary figure 2). No comparison could be performed for the SNP associated with the 6MWT (rs1108983), as all healthy individuals had the same genotype (A/A). Additionally, there were no differences in allelic nor genotypic frequencies between people with COPD and healthy individuals for the significant SNPs.

### Replication

#### 1. 6MWT and CAAT

Individuals from EARLYCOPD cohort were predominantly male (78%), presented an age of 62 [50, 71] years old, a CAAT total score of 13 [8, 20] and a total distance walk in the 6MWT of 540 [473.3, 627.7] meters (Table 1). From the SNPs associated with the 6MWT in the Lab3R-ESSUA Cohort, rs7488690 demonstrated nominal significance and an effect in the same direction as in the Lab3R-ESSUA Cohort (rs7488690:A, β (SE)= -53.7 (25.8) m, P=0.04) (Figure 3A and Supplementary table 3). The SNP rs1108983, which was genome-wide significant in the discovery cohort, was not available in EARLYCOPD cohort. Regarding the SNPs found associated with CAAT total score, three SNPs in the same loci were nominally significant and with an effect in the same direction as in the Lab3R-ESSUA Cohort (top hit rs146330339:A, β (SE)= 3.6 (1.4) points, P=0.01) (Figure 3B and Supplementary table 4). The SNPs rs11747040 and rs11041680, both genome-wide significant in the discovery cohort, did not replicate in the EARLYCOPD cohort (P=0.57 and P=0.25, respectively)

#### 2. QMVC and Handgrip

We found 31 SNPs reported in the literature as associated with the QMVC, of which three SNPs demonstrated nominal significance and equal effect direction in the Lab3R-ESSUA cohort (rs11867552:T, β (SE)= -1.4 (0.5) Kg, P=0.004; rs11942832:C, β (SE)= -12.4 (4.0) Kg, P=0.038; rs12135534:G β (SE)= 1.4 (0.7) Kg, P=0.047) (Supplementary table 5). Similarly, from the 41 SNPs reported in the literature as associated with the Handgrip, four SNPs across two loci (nearest genes *PLEKHB1* and *ANGPT2*) were nominally significant in the Lab3R-ESSUA cohort and showed an effect in the same direction of the original study (top hits rs7128512:A, β (SE)= -1.7 (0.6) Kg, P=0.009; rs752045:G β (SE)= 1.6 (0.7) Kg, P=0.02) (Supplementary table 6).

## Discussion

In this study, SNPs associated with functional capacity, muscle strength and disease impact were identified. The effect of these SNPs was independent of the presence of COPD, supporting the argument that COPD-related impairments develop on top of a pre-existing genetic predisposition, thereby contributing to the heterogeneity observed in the disease.

Using the Lab3R-ESSUA cohort, a well-characterized cohort consisting of people with COPD and healthy individuals, genome-wide significantly associated SNPs were found for 6MWT (rs1108983), the 1-min STS (rs5889103), the Handgrip (rs67352743) and the CAAT (rs11747040 and rs11041680). Additionally, rs7488690 which was suggestively associated with the 6MWT, and rs146330339 which was suggestively associated with the CAAT were validated in the EARLYCOPD cohort. Finally, the SNPs rs11867552, rs11942832 and rs12135534 that have been reported as associated with the QMVC, and the SNPs rs7128512, rs752045, rs11235843 and rs6590 that have been reported as associated with the Handgrip, were replicated in the Lab3R-ESSUA cohort.

This is one of the few studies that has explored the influence of genetic background on functional capacity and muscle strength, and the first to ever investigate the genetic factors of patient-reported outcome measures, specifically CAAT^24–26^. These traits are intrinsically connected to individuals’ ability to perform daily activities and be independent^19,20,38,52^. Importantly, these traits are often treatable (e.g., pulmonary rehabilitation), and their timely identification has been extensively advocated in the literature, as their appearance frequently precedes clinical diagnosis in disease contexts (i.e., pre-COPD)^53–56^. Therefore, identifying treatable traits at an early stage allows for targeted interventions that may slow disease progression, improve functional outcomes, and enhance quality of life^53^. A treatable traits-based approach has been proposed, supporting a precision medicine framework, shifting from a symptom-based to a proactive, patient-centred management strategy, ensuring that individuals receive personalized treatments tailored to their specific impairments and risk factors^57,58^. In this sense, our study contributes by (1) reporting SNPs significantly associated with such treatable traits; (2) providing a deeper understanding of the biological mechanisms underlying these variables; and (3) proposing genomics as a potential tool for timely identifying individuals at higher risk of impairment for these variables.

In the GWAS for the 6MWT, rs1108983 reached genome-wide significance in the Lab3R-ESSUA cohort, with a decrease of 186.5 meters per G copy in the total distance walk during the test. This SNP is located downstream of *TNFSF8*, a gene responsible for the production of CD30L. CD30L is a cytokine that, upon binding to CD30, triggers a pro-inflammatory response^59,60^. A recent study has reported higher expression levels of CD30L and CD30 in people with COPD compared with healthy individuals^61^. This would suggest that CD30/CD30L might be involved in the chronic inflammation state observed in people with COPD. Chronic inflammation, specifically in the context of COPD, has already been linked to muscle wasting/dysfunction and decreased exercise tolerance^62–64^. Based on our findings, we can speculate that the G allele of rs1108983 might trigger an over-expression of *TNFSF8*, leading to increased production of CD30L. This increased production of CD30L would sustain the state of inflammation, and negatively impact the 6MWT performance. Unfortunately, rs1108983 nor any SNP in linkage was present in the EARLYCOPD cohort, precluding its validation. In addition to the significant SNP identified, rs7488690 was found suggestively associated with 6MWT in Lab3R-ESSUA cohort and validated in the EARLYCOPD cohort. The A allele showed similar frequencies in the two cohorts, but the validation cohort showed a 30-meter smaller effect compared to Lab3R-ESSUA cohort, with both cohorts having the A allele associated with a worse 6MWT performance. Rs7488690 is an intronic variant of *LOC101928387*, a non-coding RNA gene, which has not been characterized nor associated with any trait yet.

The GWAS on the 1-min STS identified one significant SNP, rs5889103. For each GTT copy of rs5889103, the number of repetitions performed in the 1-min STS increases by 4. Rs5889103 is an intergenic variant, and expression quantitative trait loci (eQTL) analysis showed no evidence that it regulates any nearby gene, precluding its biological interpretation. GSEA highlighted three main pathways associated with 1-min STS, namely processes involving glycosylation, activation/inactivation of fast Na+ channels, and extracellular matrix remodelling, all of which have been documented in the literature in the context of COPD and other pathophysiological scenarios^64–69^. Skeletal muscle homeostasis emerges as the strongest candidate for the link with the 1-min STS, as the other two processes are related to lung pathophysiology and there is a low to moderate correlation between functional capacity and lung health state^52,70–74^. Functional experiments and fine mapping would be crucial to unravel the biological function of this SNP or others putatively linked with it and related genes.

Although no SNP in the Lab3R-ESSUA cohort reached genome-wide significance for QMVC, three SNPs that have been previously reported in the literature as associated with QMVC were validated in our cohort. Of note, rs11942832 was associated with a reduction of 12.4 Kg.F in the QMVC. The three validated SNPs were mapped to *HRNBP3, CCDC149* and *PCNXL2*, though none of these genes have previously been linked to muscle strength in the literature. GSEA identified an enrichment in genes linked to Williams-Beuren syndrome, a well-known genetic disorder characterized by intellectual disability, growth delay, ophthalmologic and orthopaedic anomalies^75^. Among the orthopaedic anomalies, muscle hypotonia is commonly observed in these patients^75,76^. As such, the molecular mechanisms underlying loss of QMVC in people with COPD and healthy individuals might share a genetic background with the molecular mechanisms of muscle mass loss in Williams-Beuren syndrome.

The second measure of muscle strength explored in this study was Handgrip, a well-established strong predictor of all-cause mortality^77^. Rs67352743 achieved genome-wide significance, with a loss of 4 Kg in handgrip strength if at least one A allele was present. Rs67352743 is an intergenic variant and there was no gene whose expression was associated with it. Additionally, no independent cohort was available for replication. From the SNPs reported in the literature as associated with Handgrip, four SNPs across two loci were significant in the Lab3R-ESSUA cohort, mapped to *PLEKHB1* and *ANGPT2* genes. No link between *PLEKHB1* and Handgrip could be established based on literature search. On the other hand, *ANGPT2* has been shown to play a positive effect on muscle fibre repair, and to be associated with grip strength, specifically in high performance athletes such as powerlifters^78,79^. This is in line with our findings, where the minor allele is also associated with increased Handgrip. Two pathways emerged as enriched from the GSEA analysis, namely *L1CAM* and *CHL1* interaction pathways, both involved in neural development and with no clear connection with handgrip muscle strength^80,81^.

Finally, the last trait explored was the impact of the disease evaluated by a patient-reported outcome measure, the CAAT. Two loci (8 SNPs in chromosome 5, and 3 SNPs in chromosome 11) were identified, although neither replicated in the EARLYCOPD cohort. The loci in chromosome 5 are located in an intronic region of *CRHBP*, a key regulator of stress response, controlling the hypothalamic-pituitary adrenal axis as well as a variety of behavioural and autonomic stress responses. CRHBP binds and therefore neutralizes CRH from over activating the hypothalamic-pituitary adrenal axis^82,83^. The CAAT questionnaire is moderately correlated with anxiety and depression, where higher levels of patient-reported anxiety and depression were associated with higher levels of CAAT, meaning a higher disease burden^19,84^. In this sense, it appears reasonable to assume that the identified SNPs might trigger an inhibition of CHRBP, leaving CHR available to over activate the hypothalamic-pituitary adrenal axis and leading to higher levels of anxiety and depression. These would then be reflected in higher scores of CAAT and a higher perception of disease burden. The loci in chromosome 11 are located in an intronic region of *OR10A3*, the gene responsible for encoding an olfactory G-protein-coupled receptor (GPRC) that initiates a neuronal response upon interacting with an odorant molecule, triggering the perception of a smell^85^. Although no direct connection can be made between *OR10A3* and CAAT score, GPRCs have been linked to psychiatric disorders, including anxiety and depression^85^. This would be in line with our previous finding on the effect of *CHRBP* on CAAT, with a genetic predisposition to higher levels of anxiety and depression leading to a higher score of CAAT and therefore the feeling of a higher disease impact by individuals. This was confirmed by a significant association between both SNPs and the hospital anxiety and depression scale in our cohort (data not shown), further suggesting a potential genetic overlap between disease impact and anxiety and depression. From the suggestive SNPs identified in the Lab3R-ESSUA cohort, one locus in chromosome 9 was replicated in EARLYCOPD, with a similar effect size in both cohort (i.e., an increase of 4 points in CAAT’s total score). The locus is located in an intergenic region, precluding the interpretation of its biological effect on CAAT.

The comparison of SNP effects between people with COPD and healthy individuals confirmed that the SNPs’ associations were specific to the trait rather than to COPD. For example, although people with COPD generally performed worse on the 1-min STS, both groups showed a similar increase of 3.7 repetitions per effect allele of rs5889103 (GTT). Consequently, people with COPD carrying the GT/GT genotype experienced the biggest impairment, resulting from having COPD on top of a genetic predisposition for poorer performance in the 1-min STS. Overall, the results being independent of COPD was expected, as all GWASs analyses were adjusted for smoking, the major risk factor for COPD. Nevertheless, these SNPs modulate extrapulmonary traits that are commonly impaired in people with COPD and might be used to stratify patients with stronger need for early intervention. Moreover, we hypothesize that our findings can be transposable to other disease contexts, where these extrapulmonary traits are also identified. These treatable traits can often be effectively managed through pulmonary rehabilitation, highlighting the importance of an early identification of patients with increased risk for functional impairment, improving the efficacy of the intervention. Further validation and functional experiments are needed to assess the robustness of our findings and to elucidate on the hypotheses proposed in this study.

## Limitations

Our study has some limitations that must be acknowledged. The overall small sample size for a genetic study may affect the robustness of results. This was prominent for the 6MWT, due to the logistical constraints intrinsic to performing the test (e.g., the need for a 30-meter straight corridor), meaning that all assessments performed by a physiotherapist in domiciliary settings could not include it. To tackle this, efforts were made to gain access to independent cohorts where our findings could be replicated, which was only possible for the 6MWT and the CAAT. Therefore, the findings on our cohort for the other extrapulmonary traits could not be replicated. Alternatively, and to circumvent this limitation, we used our cohort to validate the variants already reported in the literature and invite others to do the same with our variants. Many of our findings were in intergenic regions with no eQTL data available, which precludes the biological interpretation of the findings. Additionally, the published GWAS on QMVC reported the SNPs’ effects as z-scores rather than beta coefficients, which inhibits the comparison of effect sizes between studies. As such, only effect direction could be compared for QMVC. Finally, we explored functional capacity using the 6MWT and the 1-min STS, muscle strength using QMVC and Handgrip, and impact of disease using the CAAT. It is well known, especially in the context of COPD, that the classification of impairment for a trait varies according to the variable used (e.g., a person can be impaired for the 6MWT but not be impaired for the 1-min STS, both measures of functional capacity). As such, our results must be interpreted as specific for the measure and not specific for the “trait”. Nevertheless, the variables we’ve studied are the most commonly used in the context of COPD.

## Conclusion

In this study, we identified several genetic variants associated with extrapulmonary traits such as the 6MWT, the 1-min STS, the QMVC, the Handgrip and the CAAT. These findings provide novel insights into the genetic architecture of functional impairment, muscle weakness and disease burden, all of which are critical factors, particularly in disease contexts like COPD. While some of the identified genetic associations were replicated in an independent cohort, further validation and functional studies are necessary to confirm these results and uncover the biological mechanisms involved. This research adds to the growing body of knowledge on the genetic foundations of functional capacity and muscle strength, paving the way for future studies across a range of disease contexts.

## Data Availability

The data that support the findings of this study are not publicly available due to privacy/ethical restrictions, but are available from the corresponding authors (Rui Marcalo, Alda Marques, Gabriela Moura) upon reasonable request.

