## Supplementary figure for "Genome-wide association study of extrapulmonary traits in the context of COPD"

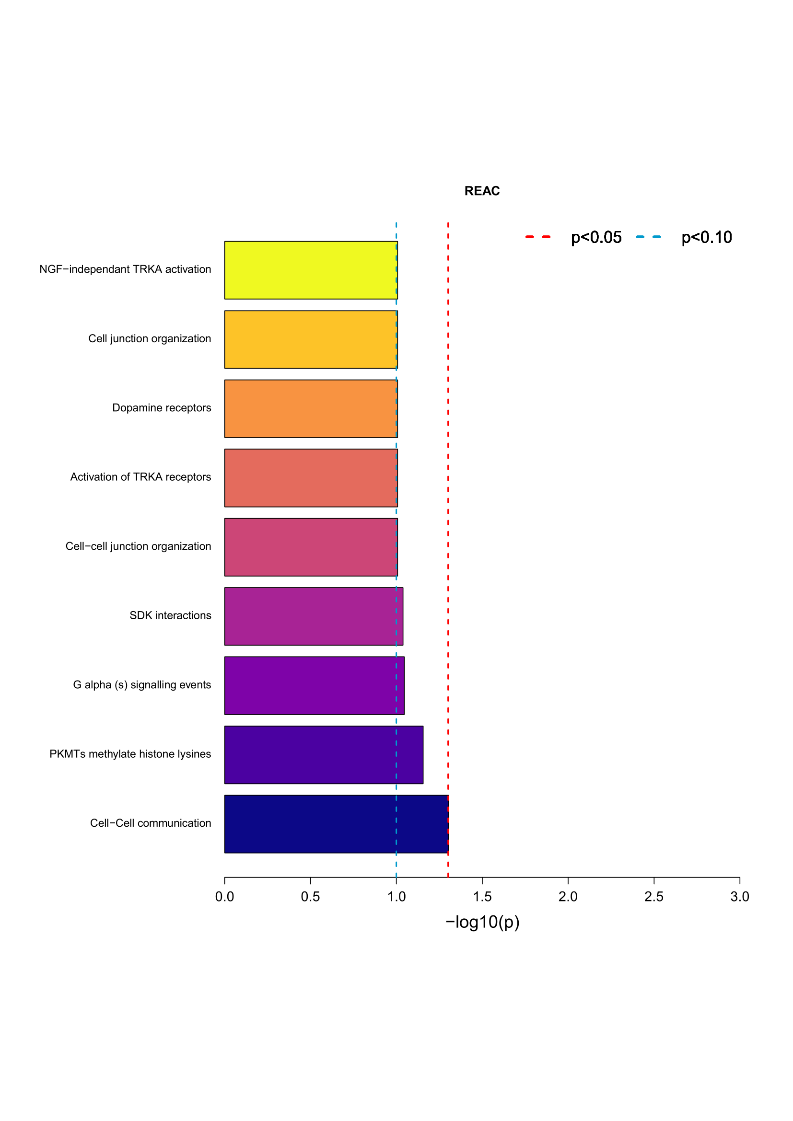

A

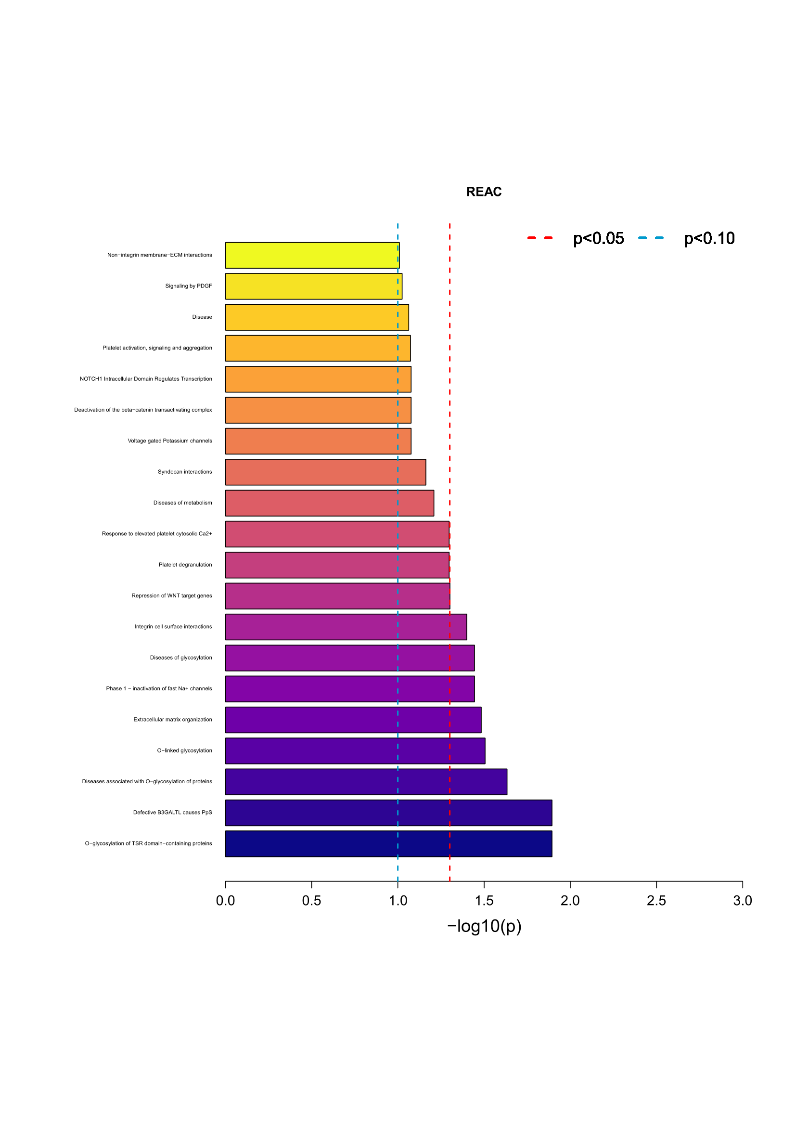

B

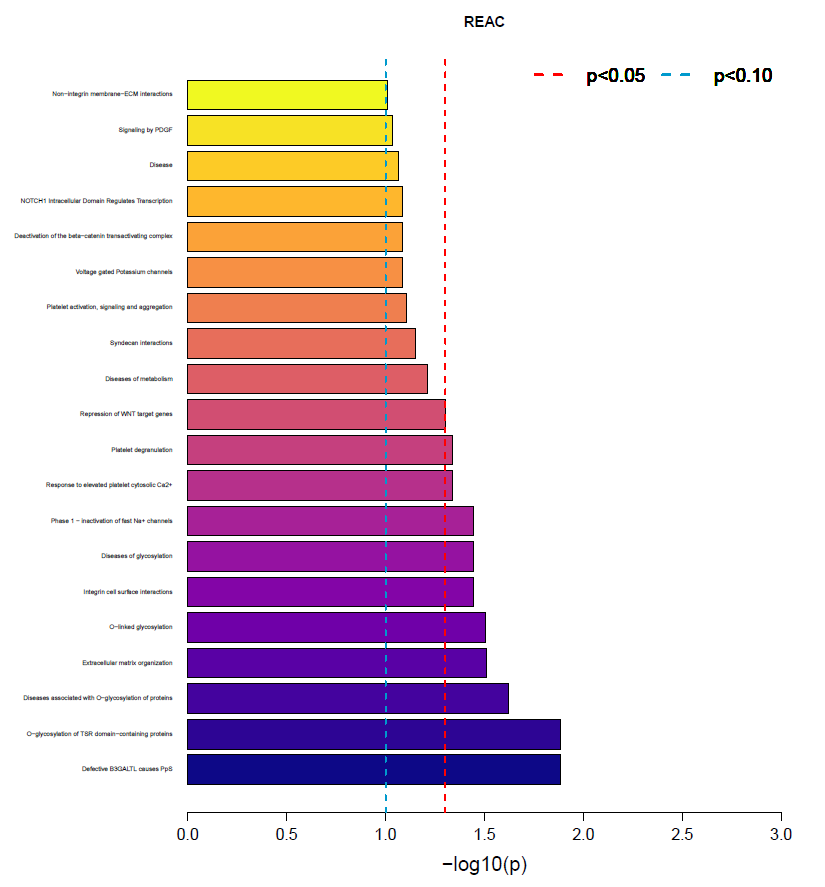

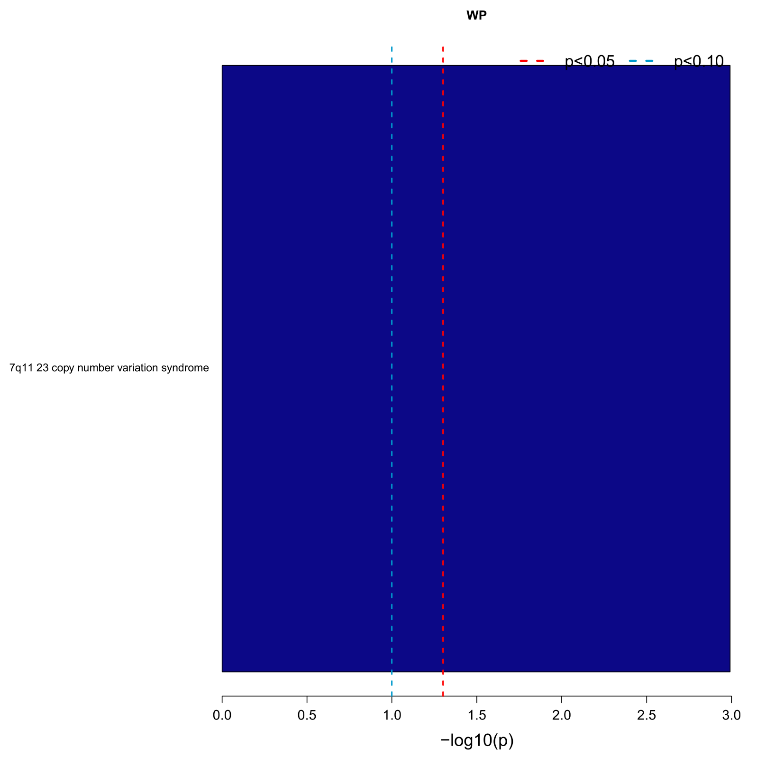

C

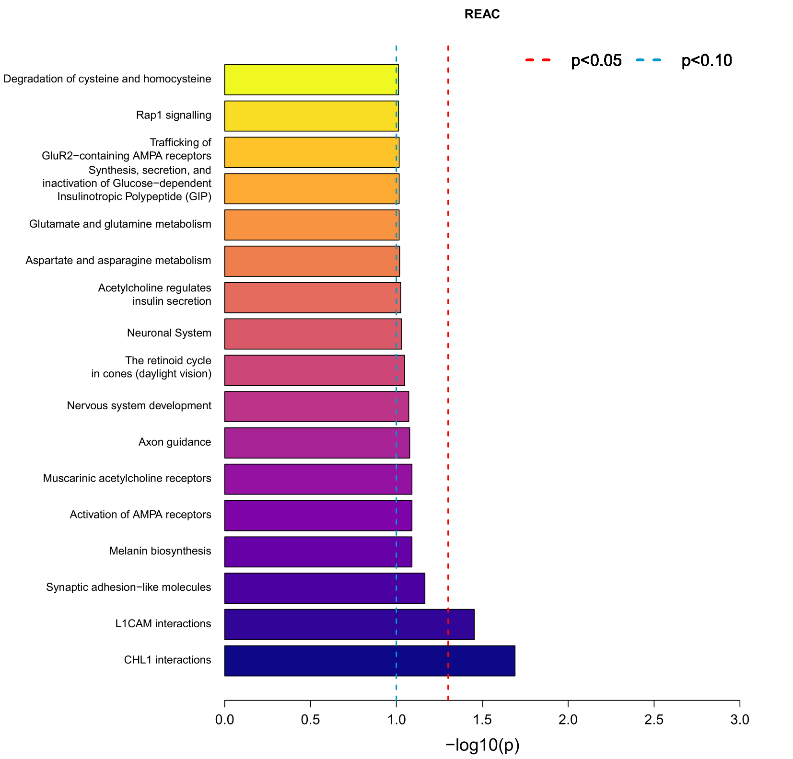

D

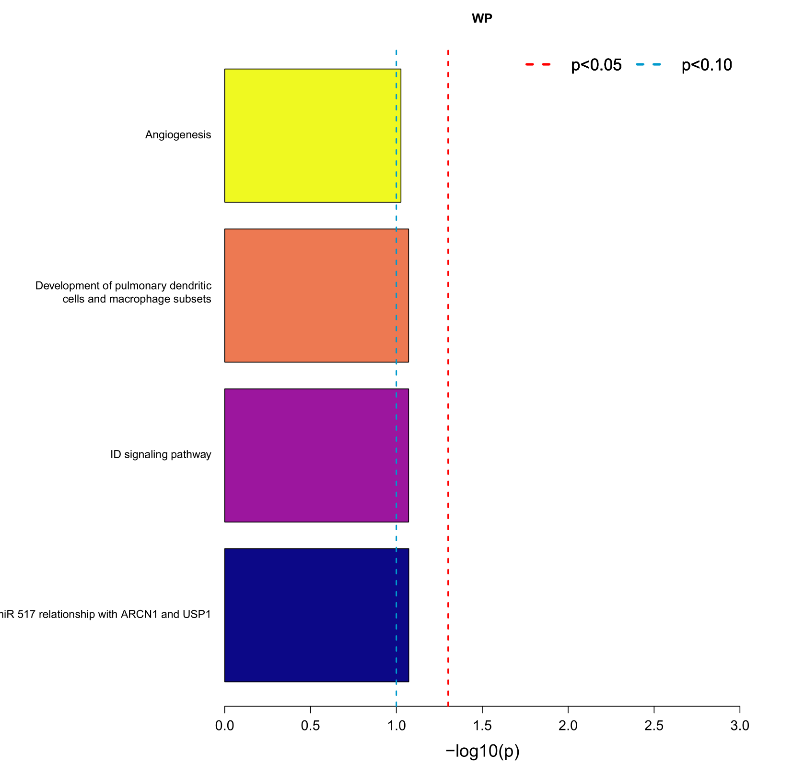

E

Supplementary figure 1. Gene-set enrichment analysis of mapped genes for (A) 6-minute walk test, (B) 1-minute sit-to-stand test, (C) quadriceps maximal voluntary contraction, (D) handgrip muscle strength and (E) chronic airways assessment test. Analysis was carried in snpXplorer online platform by submitting the list of SNPs with P<1E^-05^. REAC: reactome pathway database; WP: wikipathways database.

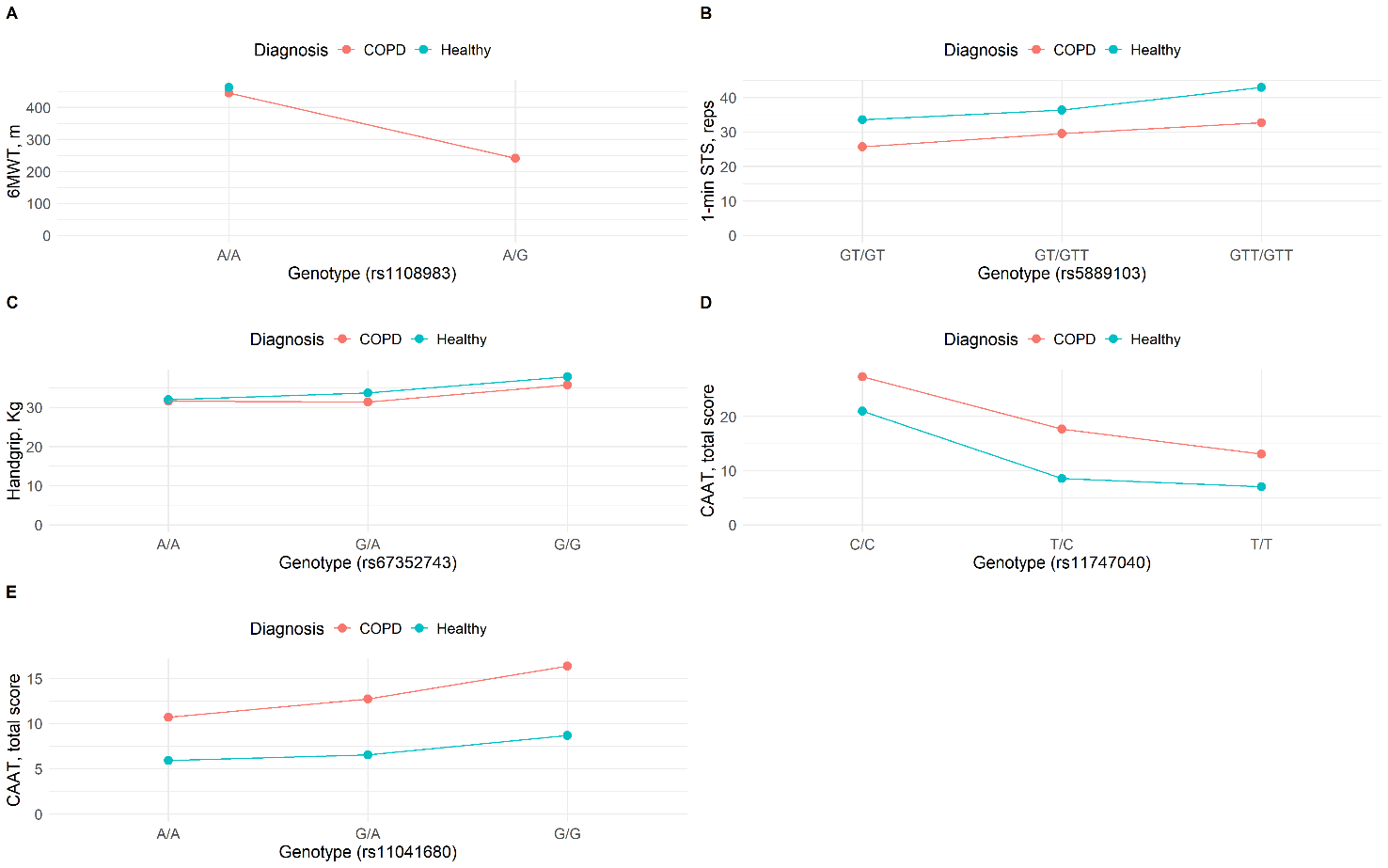

Supplementary figure 2. Mean effect of A) rs1108983 on 6-minute walk test (6MWT), B) rs5889103 on 1-minute sit-to-stand test (1-min STS), C) rs67352743 on handgrip muscle strength (Handgrip), D) rs11747040 on chronic airways assessment test (CAAT), and E) rs11041680 on CAAT, stratified by diagnosis (COPD vs healthy).
